# Predicting sensitivity to general anesthesia: Bispectral index versus Checkpoint-Decomposition Algorithm

**DOI:** 10.1101/2023.05.03.23289473

**Authors:** C. Sun, I. Constant, D. Holcman

## Abstract

Despite a large effort in EEG signal processing, classification algorithms, deep-learning approach, predicting the sensitivity to general anesthesia (GA) remains a daunting hurdle. We compare here the ability of the Bispectral Index™ (BIS™), developed more that twenty years ago to monitor the depth of anesthesia, with the real-time checkpoint-decomposition algorithm (CDA) to evaluate the patient sensitivity from the early induction phase of GA. Using EEG recorded in children anesthetised with propofol, we computed three parameters extracted from the BIS: 1-the minimum value (nadir) of the BIS, 2-the time to reach the minimum and 3-the duration spent below 40 during the first 10 minutes. Using a logistic regression procedure, we report that these parameters provide a poor prediction of sensitivity compared to the CDA, that combined the first occurrence time of iso-electric EEG traces, fraction of suppressions of the *α*-band and its first occurrence time. Finally, we correlate the BIS values with the maximum power frequency of the *α*−band, the proportion of *α*−suppressions (*α*S) and iso-electric suppressions (IES) as well as the *α* and *δ* power ratios. To conclude, the checkpoint-decomposition algorithm complements the EEG indices such as the BIS to anticipate the sensitivity to GA.

## 1 Introduction

As Machine-intelligent and classification algorithms are currently invading the clinical field, what shall we expect for their role in predicting the depth of general anesthesia (GA) compared to existing indices? We address this question by comparing a recent algorithm with a routinely used EEG based index. Real-time brain monitoring based on frontal cortical electroencephalogram (EEG) allows to evaluate neuronal activity with a few seconds delay but at a high frequency-temporal resolution. For the past twenty years, the Bispectral Index^™^ (BIS^™^), an EEG based monitor developed by the Aspect firm and now commercialised by Covidien (USA), has been used to assess the Depth of Anesthesia (DoA).

The BIS is now referred as the standard for DoA monitoring. Despite some imperfections highlighted over the years [1, 2], it remains a reliable indicator of the patient brain awareness state during the stable maintenance phase of anesthesia. The BIS provides an indicator value ranging from 0 (very deep sedation) to 100 (awake). The initial phase of induction is characterized by a BIS decrease as the hypnotic dose and sedation of the patient increase. An admissible target is to maintain a BIS in the range 40 −60 to avoid under-or over-dose and their associated deleterious side effects such as peri-operative memorisation or hemodynamic depression. The BIS values are highly correlated with the propofol concentrations in children as well as in adults. When the BIS decreases below 40, it was admitted that it is correlated with a high proportion of iso-electric suppressions (flat EEG activity periods) a signature of deep anesthesia [3, 4]. BIS is routinely used as a clinical feedback of hypnotic effects of the anesthetic agents during surgical procedure and the incidence of intraoperative awareness might be reduced, by targeting a BIS value of 50 [5].

However, the BIS is not specifically designed to prevent anesthetic drug overdose, which thus remains difficult to anticipate and could lead to postoperative complications such as delirium or cognitive disturbance. Indeed, the presence of iso-electric suppressions (IES) has been suspected to be associated with postoperative cognitive dysfunction [6, 7].

The maintenance, induction or recovery phases are not very correlated [8], thus preventing possible general predictions. Nowadays, several signal processing methods are available to analyse, or correct the EEG signal in real-time [9, 10, 11, 12]. These methods are often baesd on wavelet transform [13, 14, 15, 16, 17], independent and principal component analysis or diffusion-based classification [18, 19, 20]. Recently, we proposed transient patterns based on first passage statistical events [21] to predict the brain sensitivity to GA, such as the first occurrence time of an iso-electric suppression or a transient suppression of the *α*-rhythms (8-12 Hz) [22, 23]. The causal relation between *α*-rhythm suppressions and the first occurrence of iso-electric suppressions (IES) [22] can be used as a general predictor principle. Recently, these parameters IES, *α*− band suppression and its proportion rate increase parameter were combined in a real-time checkpoint-decomposition algorithm (CDA) to quantify the sensitivity probability to GA during maintenance from the induction phase [21]. In this manuscript, we compared three parameters directly extracted from the BIS values recorded during induction of anesthesia with propofol [4]: the minimum (nadir) of the BIS, the time to the minimum and the time spent below 40, with the values provided by the CDA. First we compare, based on logistic regression classifiers between data from both methods of assessment. In the second part, we describe the influence of propofol concentrations on different parameters of the CDA algorithm and spectral power of frequency bands. Finally, we explain how the CDA provides complementary information with respect to the BIS and thus could be used in parallel to reveal the patient sensitivity from the induction.

## 2 Materials and Methods

### 2.1 General anesthesia protocol, EEG recordings and pre-processing

General anesthesia was performed with target-controlled infusion (TCI) of propofol followed by continuous administration of remifentanil (0.25 *μ*g.*kg*^−1^.*min*^−1^). Before surgery, all subjects received premedication with oral antihistamine hydroxyzine (1 mg.*kg*^−1^). At induction, propofol was administered with an initial target concentration of 6 *μg*.*ml*^−1^. The subjects were then intubated with cuffed tracheal tube after a single dose curarization by atracurium besilate (0.5 mg.*kg*^−1^) and mechanically ventilated with an air-oxygen mixture and a breathing frequency ranging from 14 to 20 *min*^−1^ to obtain an *ET*_*CO*2_ between 30 and 35 mmHg. Propofol concentration was then kept between 2 and 6 *μg*.*ml*^−1^ according to the Schnider model for post-pubertal subjects and Kataria model for pre-pubertal subjects [24].

The decision to administer extra medication was left to the discretion of the anesthesiologist in the operating room in accordance with the institution’s standard of care protocol. The EEG monitoring was accompanied by standard monitoring, which included the Bispectral index (BIS), pulse oxygen saturation (SpO2), heart rate (CF), systolic and diastolic blood pressures (SBP, DBP), mean arterial pressure (MAP), and temperature.

EEG recordings were obtained using the Brain-Quick program stem II (Micromed, Merignac, France) with a single channel record from electrodes placed on the forehead, left and right (reference) mastoids [4]. We segmented the mechanical noise, using the EEG signal power within a sliding window of length 10 seconds with 50% overlap. A time-segment *P*_*i*_ is considered an artifact when the power within such window exceeds three times the median absolute value,

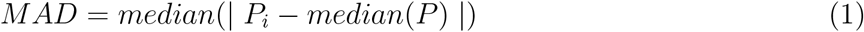

for *i* = 1, 2, .., *N*, where *N* is the number of windows. Artifact segments are then corrected using the Wavelet Quantile Normalization method [12, 25].

### 2.2 BIS recordings and estimating IES and *α*S parameters

BIS values were recorded using the BisSensor (Aspect Medical Systems) connected to the Cardiocap II monitor (Datex-GE). The recording was done continuously using Rugloop II software with a sample rate of 4 Hz. We then estimated the relevant parameters from the EEG signal and the BIS as follows:

- The time point *T*_0_ is the reference corresponding to the beginning of the induction phase after the recording started. It is usually provided in the anesthesia sheet recording.
- The first occurrence time *t*_*IES*_ of an IES and an *α*S *t*_*αS*_, as well as the slope *a* of the *α*S proportion are defined [21] with respect to the initial time point *T*_0_.
- The minimum of the BIS is the lowest value reached within the induction phase [*T*_0_, *T*_0_+10] min. We have

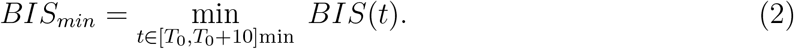
- The first peak time of the BIS is the instant where BIS reached its minimum

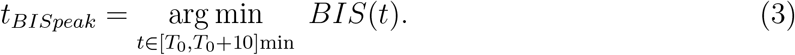
- The total time spent by the BIS under 50 during the induction is defined by

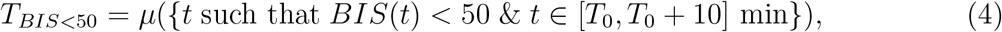

where *μ* measures the total length of interval subsets.

**Table 1:** Patients demographics, EEG and BIS statistical features.

| Variables | 50 patients |
| --- | --- |
| Age (mean ± std) | 14.5 ± 7.2 yr |
| Age (range) | 6-38 yr |
| Gender (female/male) | 36 %/64 % |
| Pubertal (pre/post) | 56 %/44 % |
| Sensitivity (IES>12s) | 44 %/56 % |
| Sensitivity ($CC_{BIS50} < 3\mu g.ml^{-1}$) | 50 %/50 % |
| Normalized first $\alpha S$ time (mean ± std) | 4.4 ± 2.2 |
| Normalized first IES time | 0.6 ± 1.0 |
| Slope of $\alpha S$ proportions | 0.8 ± 0.7 %/min |
| Normalized first BIS peak time | 1.1 ± 0.4 |
| Minimum BIS value | 20.2 ± 6.1 |
| Duration spent in BIS<50 | 7.3 ± 1.6 min |

### 2.3 BIS versus CDA parameters

To predict the patient sensitivity during the induction phase, we focused on six parameters, separated into two ensembles: three parameters are described in [21]: There are computed after induction: 1-the first occurrence time *t*_*αS*_ of an *α*−band suppression (Fig.1A, yellow segments) 2-the first occurrence time *t*_*IES*_ of an iso-electric suppression (Fig.1A, red segments). We also extracted the IES (Fig.1A, red curve) and *α*S (Fig.1A, yellow curve). 3-we estimated the slope *a* of the *α*S proportion (Fig.1A, blue dashed line), computed at 10 minutes after the hypnotic injection started *T*_0_ (Fig.1A, blue point).

**Figure 1:**
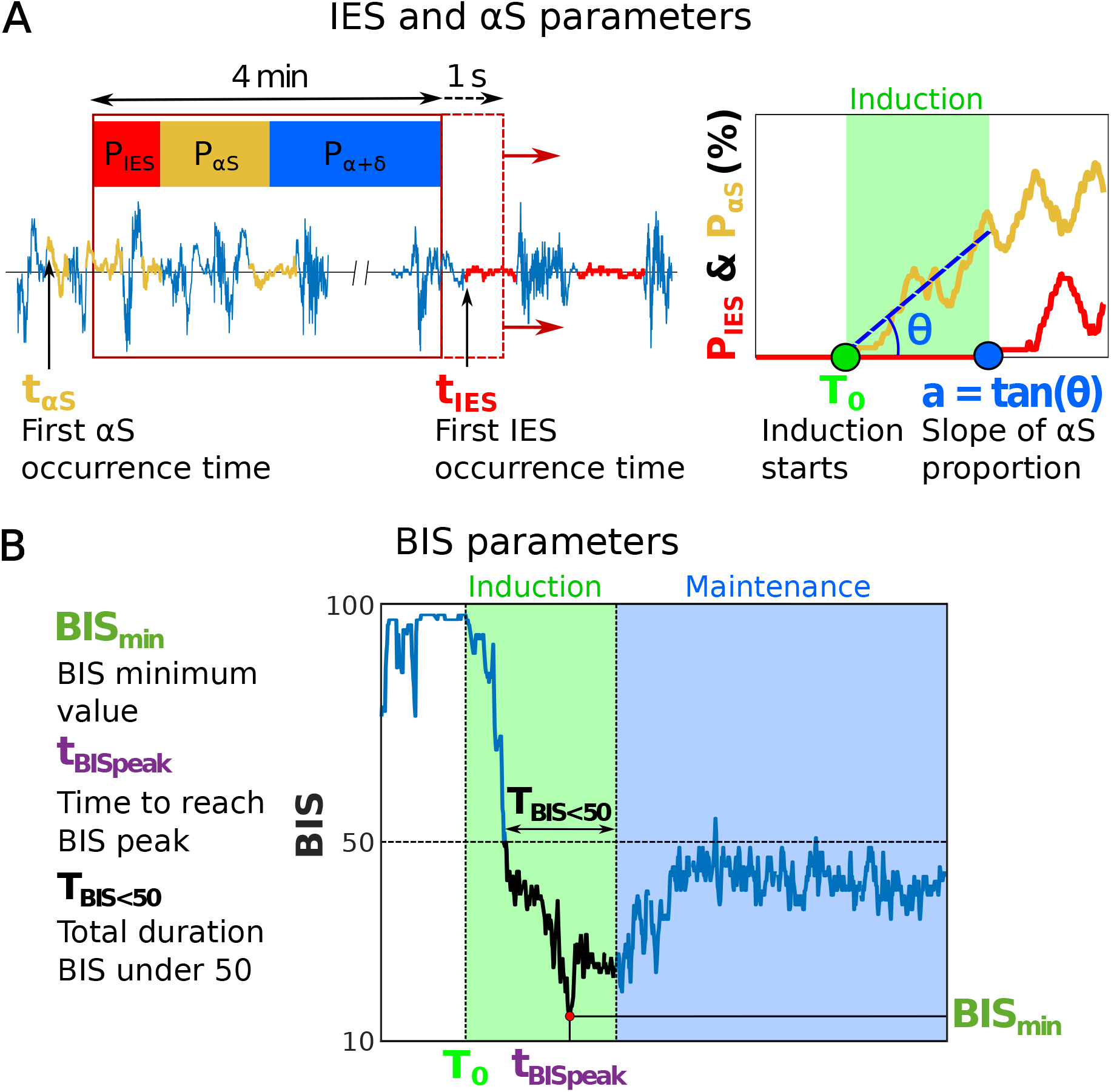
Defining IES, *α*S and BIS parameters. **(A)** Sliding window analysis used to compute the fraction of IES and *α*S. The first IES and *α*S occurrence times *t*_*IES*_ and *t*_*αS*_ are taken as the onset of the first occurrence time. The slope *a* is computed at 10 minutes after the induction phase started. **(B)** Three parameters computed from the BIS curve: the minimum value *BIS*_*min*_, the time to reach the minimum peak *t*_*BISpeak*_ and the total duration spent by BIS below 50 *T*_*BIS<*50_.

The second ensemble of three parameters is associated with the BIS. We first determined the BIS minimum value *BIS*_*min*_ to represent the maximum depth of anesthesia. Second to extract the kinetics, we use the time to reach the minimum peak *t*_*BISpeak*_ from the initial time *T*_0_ (Fig.1B green and purple). Finally, we computed the total duration during which BIS was below 50 (Fig.1B black), as a marker for the cumulative brain time response to GA. We consider that these three parameters can bring complementary information. We then decided to compare the predictive value for each set of parameters. The results are presented in the next subsection.

### 2.4 IES and *α*S segmentation

IES and *α*S were segmented following the procedure described in [21]. We first filtered the EEG signal *S*(*t*) in the range 8− 16 Hz to get the signal *S*_*α*_(*t*). We normalized *S*_*α*_(*t*) by its Root Mean Square to obtain *Ŝ*_*α*_(*t*). We computed the difference *D*(*t*) and *D*_*α*_(*t*) between the respective upper and lower envelops local maxima and minima interpolation, defining two threshold values

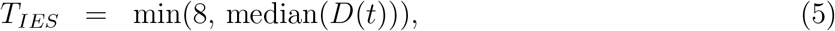

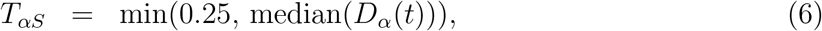

We consider an EEG segment as an iso-electric (resp. *α*) suppression when the amplitude of *Ŝ*_*α*_ is smaller than *T*_*αS*_, the amplitude of *S*(*t*) is smaller (resp. greater) than *T*_*IES*_ and it lasts at least 1 second (resp. 0.5 second).

### 2.5 Patient sensitivity labelling

We label a patient as sensitive to GA, when the EEG presented at least a minimum of 12 seconds in IES during the first 30 minutes (in the absence of any anesthesia bolus) [21]. We also define another labelling: the patient is sensitive when the propofol concentration is less than 3 *μg*.*ml*^−1^ to maintain a BIS value at 50. We separated our patients cohort into two groups: sensitive versus non sensitive.

### 2.6 Correlation measures between EEG, BIS values and anesthetic concentrations

To compare the EEG signal with the BIS values and the propofol concentrations, we developed a visual representation allowing to monitor simultaneously the time evolution of the EEG and spectrogram, which represents the time-frequency evolution of the waveform. The spectrogram is computed using Short Time Fourier Transform [26] (Fig.2A-B). We segmented the EEG waveform amplitude in IES (red) and *α*S (yellow), (Fig.2C). The proportions of IES and *α*S are computed over a sliding window of 4 minutes with a one second overlap. In parallel, we tracked the BIS values (Fig.2D) and the target propofol *C*_*t*_, the effect site *C*_*e*_ and the plasmatic *C*_*m*_ concentrations (Fig.2E).

**Figure 2:**
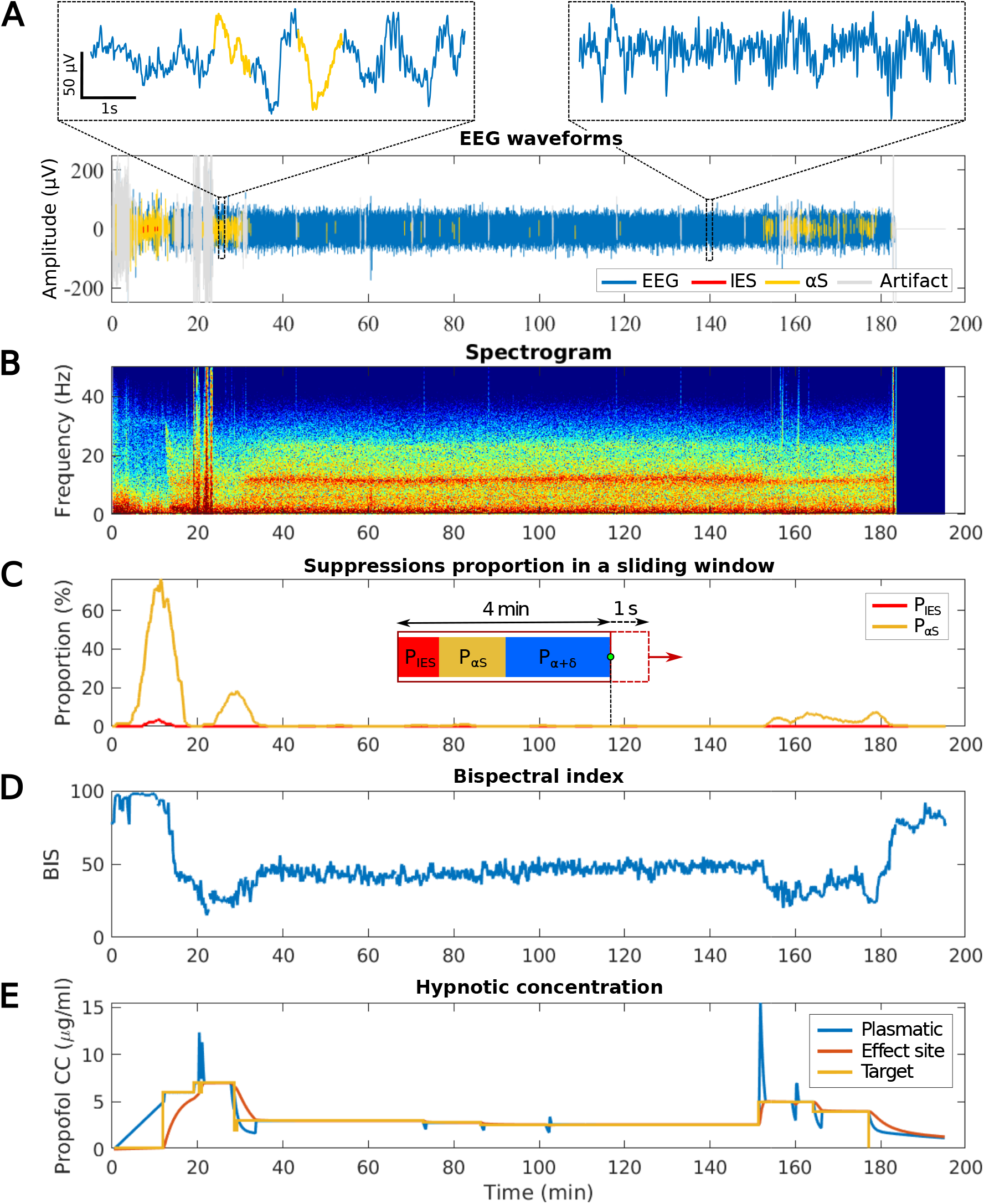
Simultaneous tracking of EEG, spectrogram, Bispectral index and hypnotic concentrations over the entire GA. **(A)** EEG signal (blue) during propofol induced GA segmented in iso-electric suppressions (IES) (red), alpha-suppressions (*α*S) (yellow) and artifacts (grey). **(B)** Spectrogram (Time-frequency) representation of the signal shown in (A). **(C)** Proportion of IES (red) and *α*S (yellow) evaluated in a 4 minutes sliding window and 1 second overlap. **(D)** BIS derived from the EEG signal, evaluating the depth of anesthesia. **(E)** Concentration of anesthetic agent (propofol) injected: the target concentration (yellow) is adjusted by hands, the plasmatic (blue) and effect site (orange) concentrations are computed according to pharmacokinetics and pharmacodynamics (PK/PD) models.

We describe now how we generated binary vectors for correlating BIS with IES and *α*S. For that goal, we generated two binary vectors *A* = (*a*_1_, ..) and *B* = (*b*_1_, …), where *a*_1_, *b*_1_ ∈ {0, 1}. We first partitioned the EEG time series in discrete time points (*t*1, *t*2, …). When the alpha-band is present (resp. absent) during the consecutive time point *t*_*k*_, ..*t*_*k*+*q*_, we set *a*_*k*_ = 1, ..*a*_*k*+*q*_ = 1 (resp. *a*_*k*_ = 0, …, *a*_*k*+*q*_ = 0). We then construct the following vectors:

- 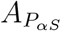 has coordinates 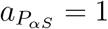 when the proportions of *α*S exceeds 5% in a sliding window and zero otherwise.
- 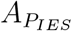 has coordinates 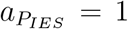 when the proportions of IES exceeds 5% in a sliding window and zero otherwise.
- 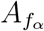 has component 1 when the maximum power frequency inside the band (6-30 Hz) is located inside the *α*−band (8-14 Hz). We apply the same procedure for constructing the vector *B* associated to the presence or absence of the BIS in the following cases:
- 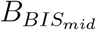: 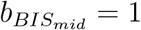 when the BIS is between 40 and 60 and zero otherwise.
- 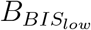: 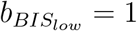 when the BIS is less than 40 and zero otherwise.
- 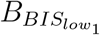 : 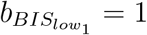 when the BIS is between 30 and 40 and zero otherwise.
- 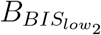: 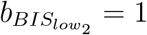 when the BIS is less than 30 and zero otherwise.

To assess the relation between these binary vectors, we use the cosine similarity formula defined by the relation

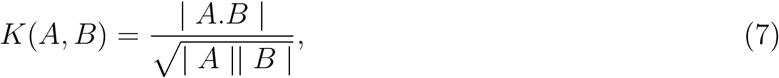

where A and B are the two vectors and |*A*| is the number of 1 in A and *A*.*B* is the scalar product. The similarity *K* measures the presence of element of A and B at the same position: when the two vectors are made of 1 all the time, *K* = 1. When the coordinates of *A* and *B* are never one at the same position, then *K* = 0. Note that the computation of the cosine similarity is independent of the partition in time.

### 2.7 Determining the patient sensitivity probability *P* (*S* = 1)

To evaluate the patient sensitivity, we first apply a logarithm transformation to the normalized first *α*S and IES occurrence times. We then quantified the predictive value of the six parameters using the Random Sub-Sampling Cross-Validation method [27] which consists of generating multiple random data splits by a fixed ratio of 70% for training and 30% for testing. In each sampling, by considering each combination of the three parameters, we trained and tested a logistic regression model [28] for the two sensitivity criteria.

Specially, we estimated the patient sensitivity from the two sets of variables: one coming from the BIS and the other from CDA. The overall patient sensitivity is computed as the output of the logistic regression classifier with the parameters collected during the induction phase of GA. The conditional probability for the sensitivity *S* to take either 0 (non sensitive) or 1 (sensitive) is defined by

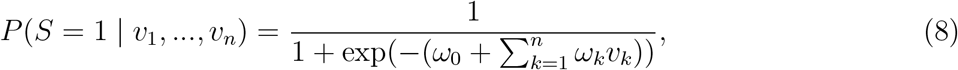

where the variables {*v*_1_, …, *v*_*n*_} are the parameters {*a*, 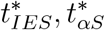, *BIS*_*min*_, *t*_*peak*_, *T*_*BIS<*50_*}* and the coefficients (*ω*_0_, …, *ω*_*n*_) are estimated from the logistic regression optimization procedure, where the cohort is split into 30% and 70%.

### 2.8 General statement about data processing

This prospective observational randomized study included children patients receiving a scheduled elective procedure requiring general anesthesia. We used a similar cohort as the one described in [2]. For each patient, two or more plateaus of 10 minutes duration were applied with a fixed concentration: one or more random concentrations and one for which the BIS value was maintained at 50. This study was approved by the Ethics Committee CPP Saint Antoine, Paris, France (Approval number: 04605). Written and informed consent was obtained from children and their parents. The protocol has been therefore performed in accordance with the ethical standards laid down in 1964 declaration of Helsinki and its later amendments. A group of 68 patients ranging from 6 to 38 years old, ASA 1 or 2, included for middle ear surgery, was used in this study. Among the patients cohort, 50 had complete data (BIS curve, concentrations data, readable spectrogram) and we discarded the remaining ones from the study. Data were collected between 2005 and 2006. The trial was later on registered and completed on September 2016 (ClinicalTrials.gov ID: NCTO2893904). We used MATLAB R2021a software to perform the statistical analysis. The significance level used in this study was *α*=0.05. The values were expressed in mean± standard deviation. Variables were compared using Student’s t-test or Wilcoxon-Mann-Whitney test as appropriate.

## 3 Results

### 3.1 BIS parameters do not predict sensitivity to general anesthesia

To evaluate the patient sensitivity, we compared several combinations of the three parameters (see method) and plotted the AUC distribution from 50 sampling in Fig.3A. Additionally, we plotted the mean ROC-AUC curves (Fig.3B-C) and we found here that the univariate parameter *t*_*IES*_ (first IES occurrence time) was the best predictive score with an *AUC* = 0.84 ± 0.10 when predicting the IES sensitivity and an *AUC* = 0.72 ± 0.11 for the concentration sensitivity. However, among the three BIS parameters, the highest mean AUC obtained 0.53 ± 12 was obtained with the BIS minimum value *BIS*_*min*_, which represents a 21% mean gap with the best CDA parameter. The remaining two BIS parameters had an AUC less than 0.50, which by definition is worse than a perfectly random classifier. We conclude that the three BIS parameters cannot be used to evaluate the sensitivity to GA in comparison with the ones defined for the CDA.

**Figure 3:**
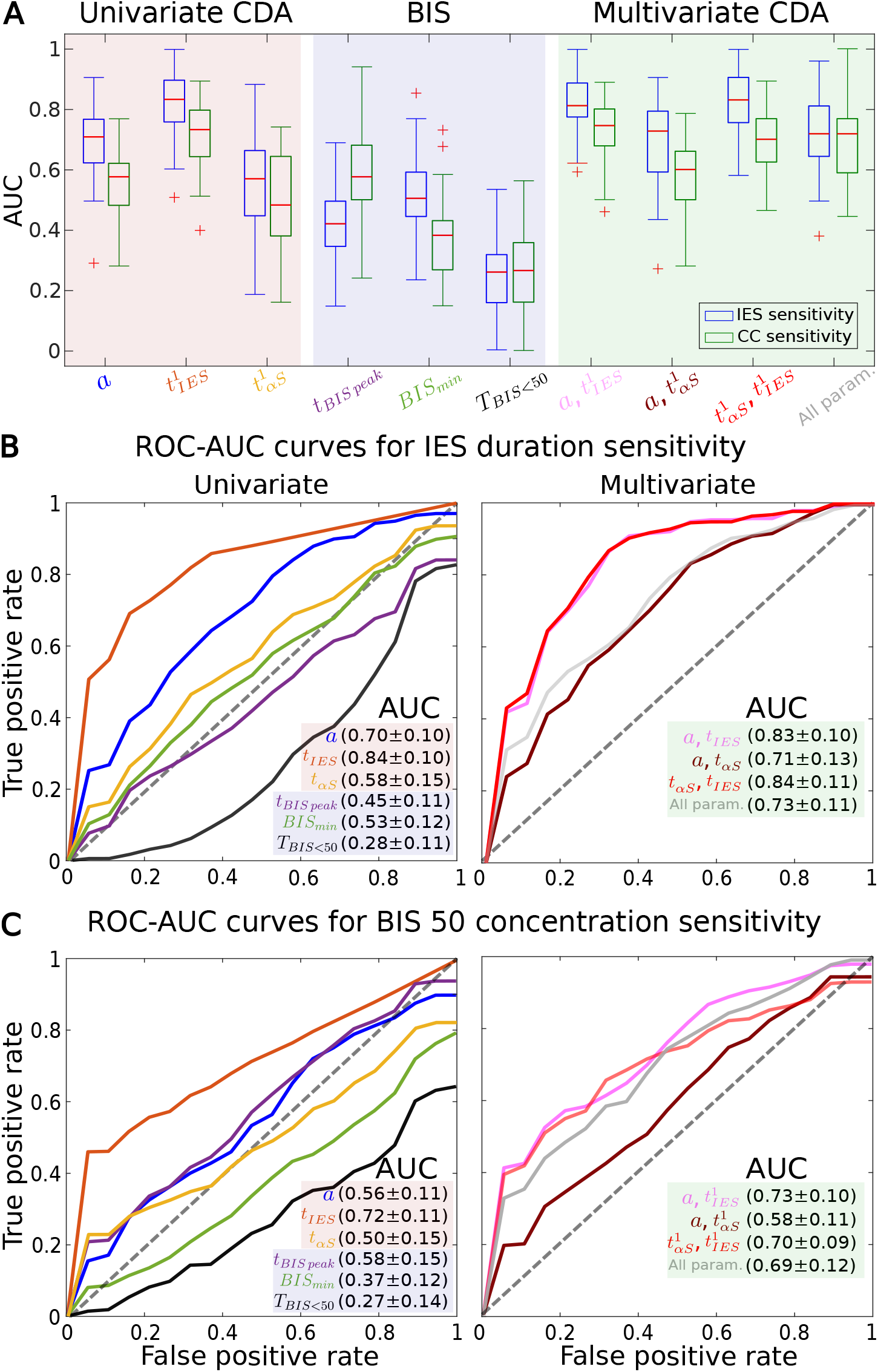
Comparing the predictions between IES and concentration based sensitivities using IES, *α*S and BIS parameters. **(A)** AUC distribution of the univariate checkpoint decomposition algorithm (CDA) (red background), BIS (blue) and multivariate CDA (green) parameters. The logistic regression classifier uses 50 random samplings of the patients cohort. **(B)** Mean ROC-AUC curves to predict IES sensitivity: slope (blue), first IES (orange), first *α*S (yellow), first to BIS minimum peak (purple), BIS minimum value (green) and total duration the BIS remained below 50 (black), slope and first IES (pink), slope and first *α*S (brown), first IES and *α*S (red) and all three parameters (dark gray) (multivariate). **(C)** Same as (B) but for the concentration sensitivity.

### 3.2 Comparing two criteria of sensitivity

We compare the two criteria of sensitivity (one based on a total time spent in iso-electric suppression that exceeds 12 seconds and second based on hypnotic concentration at BIS value 50) and found that the two criteria overlap for only 70% of cases as shown in the matrix of Fig.4A. Thus for 30% of patients, BIS50 sensitivity is not associated to the IES sensitivity. Furthermore, we investigated the correlation between patient age and the total time spent in suppressions for the two sensitivity criteria Fig.4B-C: for the IES sensitive patients, we found that the Spearman correlation between age and total spent in IES (resp. *α*S) is *ρ* = 0.10, p-value= 0.12 (resp. *ρ* = −0.23, p-value= 0.32). Similarly, for BIS50 sensitivity, we found a Spearman correlation of *ρ* = 0.11, p-value= 0.62 (resp. *ρ* = 0.00, p-value= 0.99) between age and total IES (resp. *α*S) duration. We conclude that there was no correlation between patient age and the total time spent in IES (resp. *α*S) for both sensitivity criteria.

**Figure 4:**
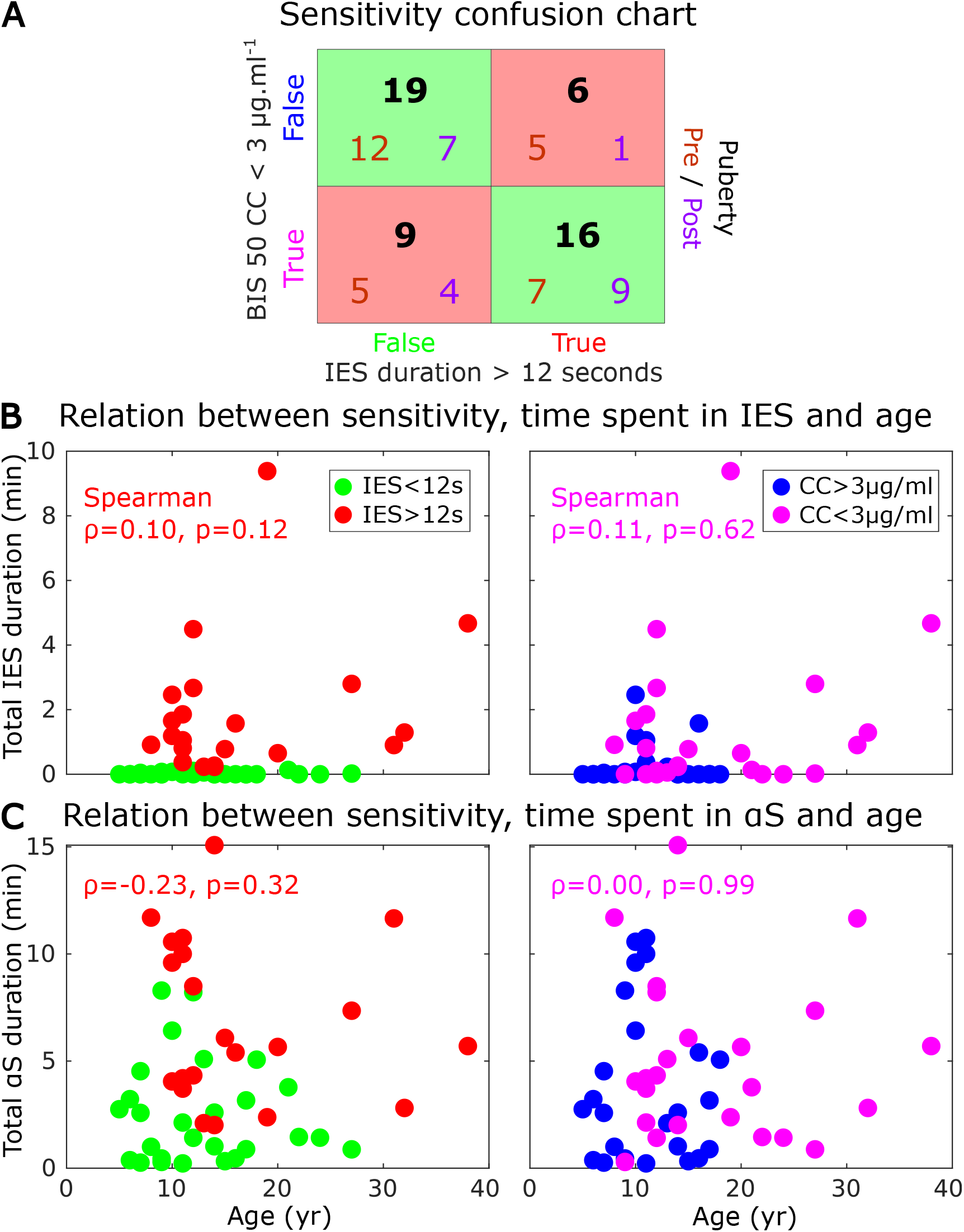
Age, total duration spent in suppressions and sensitivity. **(A)** Confusion chart for two criteria of sensitivity: 1-total IES duration exceeding 12 seconds and 2-concentration (CC) for BIS equals 50 less than 3 *μg*.*ml*^−1^. We differentiated Pre (red) versus post (purple) puberty. Most of the false negative and positive occurs for the pre-puberty group. **(B)** Scatter plots between the age and the total time spent in IES grouped by IES (left) and concentration (right) sensitivity. **(C)** Scatter plots between the age and the total time spent in *α*S grouped by IES (left) and concentration (right) sensitivity.

### 3.3 *α*−band appearance is associated with BIS values between 40 and 60

To better correlate the ranges over which the BIS values evolve and the presence of IES and/or *α* −suppressions, we use the cosine similarity (Eq.(7), see Material and Methods). We shall compute this index between the BIS and the proportions of IES, *α*S, power of the bands *α* and *δ*, as well as the maximum power frequency curve. The binarization procedure is described in the subsection 2.6. We found that the cosine similarity between the BIS value *BIS*_*mid*_ between 40 and 60 and the maximum power frequency *f*_*α*_ is 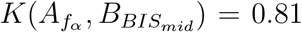 [0.71, 0.88] (Median [IQR]). However, when the BIS is less than 30, as measured by the vector 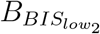, and the proportion of IES (resp. *α*S) exceeds 5% (summarized in the vector 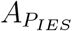, the similarity is 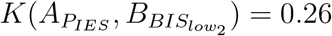) = 0.26 [0.14, 0.54] (resp. 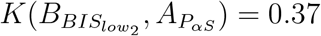 [0.21, 0.53]). For BIS value between 30 and 40 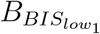, the similarity with the IES (resp. *α*S) proportion is 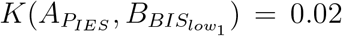 [0, 0.09] (resp. 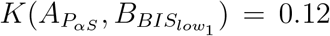 [0.06, 0.31]) (Fig.5A). At this stage, we conclude that the BIS is associated with the presence of a maximum frequency *α*− band and barely with the IES and *α*S proportions.

**Figure 5:**
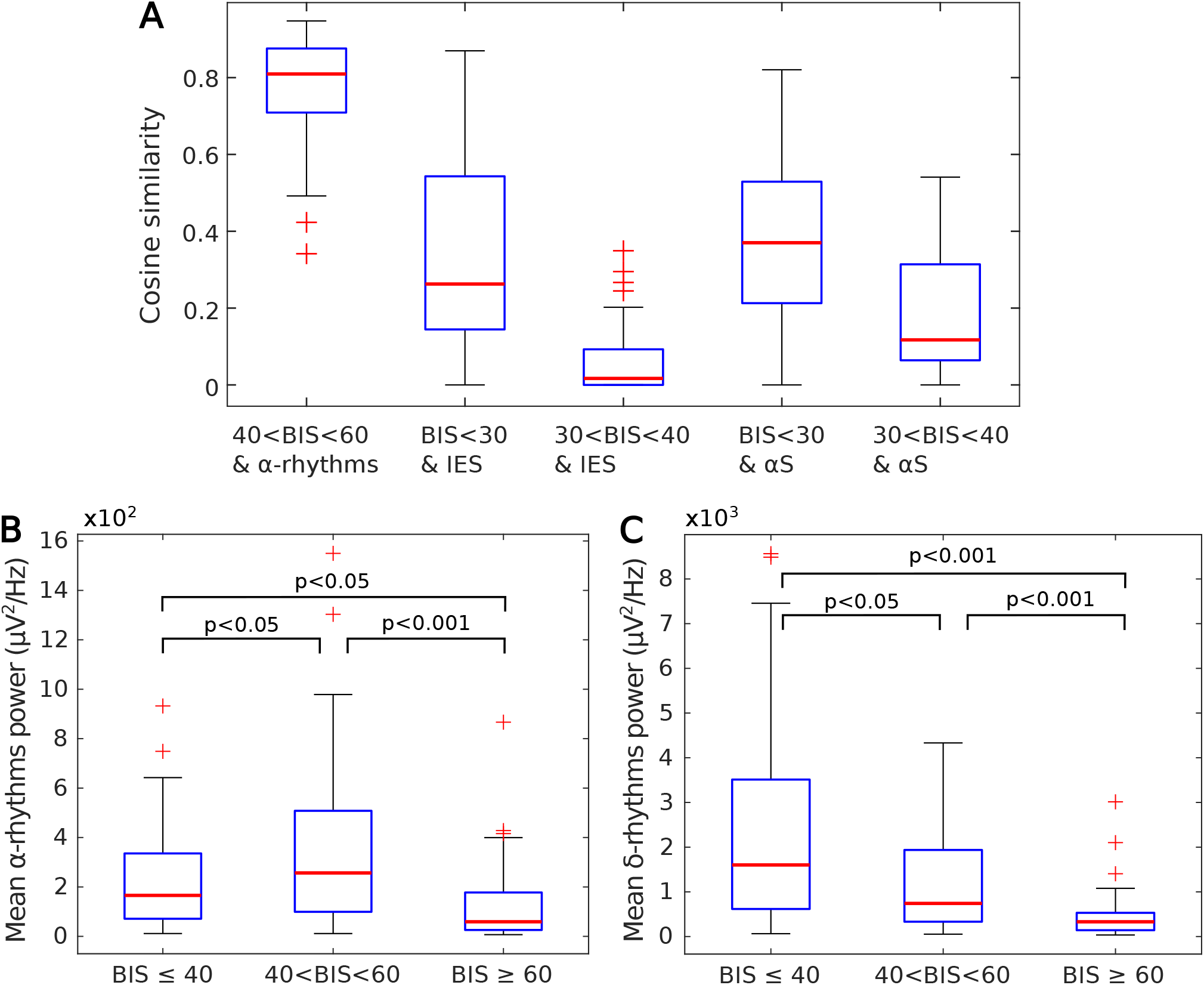
Comparing BIS range values with bands suppressions and power. **(A)** Cosine similarity distributions between two binary sequences obtained from the following conditions: 1) BIS values in the range 40 and 60 and maximum power frequency within the *α*-oscillations. 2) BIS value below 40 and IES are detected. 3) BIS under 40 and *α*S are detected. **(B)** Distribution of the mean *α*-band power and three BIS ranges values: BIS<40, 40<BIS<60 and BIS>60. **(C)** Mean *δ*-band power distribution and three BIS ranges values as in (B).

Furthermore, we decided to explore how the *α* −rhythms mean power is distributed with respect to three ranges of BIS values: 1) *BIS* 40, the mean power is 166 [71, 336] *μV* ^2^*/Hz*; 2) 40 ≤*BIS*≤ 60, we have 257 [99, 508] *μV* ^2^*/Hz*; 3) 60 ≥*BIS*, we have 59 [26, 178] *μV* ^2^*/Hz*. These distributions are significantly different: between 2) and 3) we found *p* < 0.001, 1) and 3) *p* < 0.05, 1) and 2) *p* < 0.05, as shown in Fig.5B. Similarly, we reported that the *δ*− rhythm mean power decreases as the value increases in the following three ranges: 1) 1602 [617, 3512] *μV* ^2^*/Hz*; 2) 741 [332, 1937] *μV* ^2^*/Hz*; 3) 331 [142, 531] *μV* ^2^*/Hz*. Again, these distributions are significantly different: between 1) and 2), we obtained *p* < 0.05, 2) and 3) *p* < 0.001, 1) and 3) *p* < 0.001 (Fig.5C). To conclude, the mean power of the *α* and *δ*− rhythms behave differently depending on the three BIS value intervals. We statistically confirmed that a BIS between 40 and 60 is associated with the presence of the *α* −band. Finally, when the BIS decreases, the power of EEG signal is concentrated on slower frequencies.

## 4 Discussion

In the present manuscript, we compared the ability to anticipate the appearance a large amount of IES between the Bispectral Index^™^ (BIS^™^) and the Checkpoint-Decomposition Algorithm (CDA) introduced in [21]. Using a logistic regression approach, this algorithm allows to predict the sensitivity to GA directly from the induction phase by extracting and combining three parameters: the first occurrence time of an iso-electric suppression, the first occurrence time of a transient suppression of the *α*-rhythms (8-12 Hz) and the increasing rate for the proportion of *α*− band suppression. These parameters quantify key events in monitoring brain sensitivity and allow to detect the trends to generate too many IES, a marker of brain sensitivity to GA. Although these parameters were previously validated, there was no comparison with respect to other methods such as the routinely used BIS, commercializing by Medtronic.

Following the step of proposing a real-time method that could be used to evaluate the sensitivity and the dose adaptation and administration, we compared here the CDA with BIS parameters (minimum, time to reach the minimum and the time spent below 50) during the first 10 minutes. Previously, the general consensus was to use a low minimum of the BIS (nadir) [2] as a marker of sensitivity. We showed here that this marker is not efficient.

Furthermore, we reported that the CDA was able to predict the sensitivity to anesthesia with propofol in a population of children and young adults. Among the three parameters we extracted from the BIS, we reported that the minimum value had a better AUC compared to the other two. This was in agreement with previous study [3] suggesting that the BIS values under 40 decreases proportionally as the ratio of IES increased. A low minimum BIS defines the presence of IES and thus is related to sensitivity. We conclude that the CDA [21] can identify the trends to generate IES, which is not the case of BIS procedure that provides an alert when IES are already present.

We highlighted here the relationship between anesthetic concentrations and the spectrogram behavior of the alpha-band suppression and power (Fig.5A-B). In particular, while we confirm that a BIS between 40 and 60 is strongly correlated with the presence of an alpha band, BIS under 30 is mostly associated with IES and *α*S. When propofol concentration increased, the power of the *α* band decreased, while the delta power becomes dominant (Fig.5C-F).

The large inter-individual variability brain responses to hypnotic such as sevoflurane or propofol calls for a general approach to identify the sensitivity trends. Pharmacological model kinetics or dynamics approximation provide time scale but do not reveal the exact sensitivity, confirming the general interest for the present type of real-time algorithms, that can connect conceptualized real-time engineering methods with clinical needs.

### 4.1 Sensitivity versus dosage

Avoiding anesthetic overdose is relevant especially to avoid damaging the brain of young children [29]. This task requires quantifying brain sensitivity that could be possible using EEG. Similarly in the brain of elderly people, suppression of the neuronal cortical activity appears with IES segments [30], occurring in patients with damaged cortical circuits or cognitive deficit. The high number of correlations between IES and depressed EEG and postoperative complications, alteration of behavior confirms the general interest to individualize anesthesia and thus to restrict the therapeutic range, as we can now recognize that anesthesia overdose is actually more predominant than expected.

As show in the present study, the present CDA algorithm could be used in parallel with the BIS to define the sensitivity of a patient from the induction phase of GA. It would have the advantage to alert the practitioner about the patient sensitivity. It remains premature at this stage to define how sensitivity could be translated into an optimal dose regulation. Each patient has indeed an intrinsic excitation-inhibition balance of neuronal circuit that evolves with age and controls the thalamo-cortical loop [31]. Transient EEG suppressions are a response to pharmacological brain perturbation. Whether sensitivity could be encoded in the suppression time patterns or not still remains difficult to comprehend.

### 4.2 Population specificity

The children population seems to be quite robust to anesthesia and thus monitoring their sensitivity is necessarily similar to adult. The sensitivity to GA can be due the specific pharmacokinetics, high metabolic rate, resulting *in fine* probably in higher doses in children compared to adults [2]. This concept probably led to many overdoses, a situation that could be studied based on the present CDA and could be extended to TCI propofol or the anesthetic remifentanil. We conclude that the present CD-algorithm could be used for clinical evaluation on different cohorts from children to elderly and characterize the difference of sensitivity generated by sevoflurane versus propofol. It would also be interesting to evaluate the algorithm on very young children, anesthetised with propofol and in particular check whether it could be possible to avoid burst, due to their cortical sensitivity, especially before one year old. There are few alpha band before 7 to 8 months old, suggesting that other form of oscillations could be relevant to detect the sensitivity to GA.

## Data Availability

Data available upon reasonable request

## Abbreviations

EEG: ElectroEncephaloGram;
GA: General Anesthesia;
TCI: Target Controlled Infusion;
*α*S: Alpha-Suppression;
IES: Iso-Electric Suppression;
BIS: Bispectral index

## Acknowledgments

D.H. research is supported by “Fondation pour la Recherche Médicale” (Postdoctorat en France - SPF201909009284), an ANR grant NEUC-0001 and the European Research Council (ERC) under the European Union’s Horizon 2020 research and innovation programme (grant agreement No 882673).

## Competing interests

The Authors declare no competing financial interests.

## Data availability

The datasets generated and analysis in the current study are available from the corresponding authors upon reasonable request.

## Notes

### Competing Interest Statement

The authors have declared no competing interest.

### Funding Statement

This study did not receive any funding

### Author Declarations

For each patient, two or more plateaus of 10 minutes duration were applied with a fixed concentration: one or more random concentrations and one for which the BIS value was maintained at 50. This study was approved by the Ethics Committee CPP Saint Antoine, Paris, France (Approval number: 04605). Written and informed consent was obtained from children and their parents. The protocol has been therefore performed in accordance with the ethical standards laid down in 1964 declaration of Helsinki and its later amendments.

